# Effectiveness and Safety of Bempedoic Acid Across Clinically Relevant Subgroups: Insights from the CLEAR Taiwan Study

**DOI:** 10.64898/2026.06.16.26355838

**Authors:** Yen-Wen Wu, Dong-Yi Chen, Chih-Sheng Chu, Yi-Yao Chang, Bing-Hsiean Tzeng, Tien-Chi Huang, Heng-Hsu Lin, Wen-Po Chuang, Chi-Cheng Huang, Jih-Kai Yeh, Chun-Yuan Chu, Ming-Yun Ho, Ching-Ya Huang, Wei-Chen Yang, I-Chang Hsieh, Tsung-Hsien Lin

**Affiliations:** Division of Cardiology, Cardiovascular Medical Center, and Department of Nuclear Medicine, Far Eastern Memorial Hospital, New Taipei City, Taiwan; School of Medicine, National Yang Ming Chiao Tung University, Taipei, Taiwan; Graduate Institute of Medicine, Yuan Ze University, Taoyuan, Taiwan; Department of Cardiology, Chang Gung Memorial Hospital, Linkou Branch, Taoyuan, Taiwan; Division of Cardiology, Department of Internal Medicine, Kaohsiung Medical University Hospital, Kaohsiung, Taiwan; Department of Internal Medicine, School of Medicine, College of Medicine, Kaohsiung Medical University, Kaohsiung, Taiwan; Medical Affairs, Daiichi Sankyo Taiwan Ltd., Taipei, Taiwan; Graduate Institute of Clinical Medicine, College of Medicine, Kaohsiung Medical University, Kaohsiung, Taiwan

**Keywords:** bempedoic acid, low-density lipoprotein cholesterol, pragmatic study, subgroup, Taiwan

## Abstract

**Background:** Despite available lipid-lowering therapies (LLT), many patients fail to achieve low-density lipoprotein cholesterol (LDL-C) targets. This gap persists across clinically relevant subgroups. Bempedoic acid has demonstrated effective LDL-C lowering with a favorable safety profile in the CLEAR Taiwan study; however, its effects across subgroups in Asian populations remains limited.

**Methods:** The phase IV CLEAR Taiwan study (NCT06925100) enrolled patients with inadequately controlled hypercholesterolemia who received bempedoic acid for 12 weeks in addition to background LLT. This analysis evaluated changes in lipid parameters, high-sensitivity C-reactive protein (hsCRP), and safety outcomes in clinically relevant subgroups, including cardiovascular risk, diabetes, age, statin tolerance, and sex.

**Results:** A total of 180 patients were included. Bempedoic acid achieved significant LDL-C reductions in all subgroups. Numerically greater LDL-C reductions were observed in primary prevention, statin-intolerant, younger (< 65 years), and female patients, while comparable reductions were observed across diabetes status. Reductions in non-high-density lipoprotein cholesterol, total cholesterol, and apolipoprotein B were consistent with LDL-C findings. Significant decreases in hsCRP were observed in all subgroups, with numerically greater reductions in patients aged < 65 years and those without diabetes. Bempedoic acid was well tolerated, with a low incidence of adverse events and no new safety signals identified. Changes in liver enzymes, renal function, and uric acid were minimal within subgroups.

**Conclusion:** Subgroup analyses from the CLEAR Taiwan study demonstrate consistent efficacy and safety of bempedoic acid across clinically relevant subgroups and support its use as a flexible option to address residual gaps in lipid management.

**Registration:** URL: https://clinicaltrials.gov/study/NCT06925100; Unique identifier: NCT06925100

**Clinical Perspective:** *What Is New?:*

- This subgroup analyses of CLEAR Taiwan study provides the real-world evidence demonstrating that bempedoic acid leads to clinically meaningful reductions in LDL-C, lipid parameters, and hsCRP across clinically relevant subgroups, including cardiovascular risk, diabetes, age, statin tolerance, and sex. Bempedoic acid was well tolerated, with a low incidence of adverse events and no new safety signals identified in subgroups.
- LDL-C reductions appeared to be numerically greater in primary prevention, statin-intolerant, younger (< 65 years), and female patients, which may be partly explained by differences in baseline LDL-C level and lipid-lowering therapy intensity.

*What Are the Clinical Implications?:*

- Bempedoic acid is effective and safe across a range of clinically relevant patient populations, including those commonly encountered in routine practice who present therapeutic challenges. These results support the role of bempedoic acid as a flexible treatment option to address residual gaps in lipid management and may help inform more individualized lipid-lowering strategies in clinical practice.

## INTRODUCTION

Elevated low-density lipoprotein cholesterol (LDL-C) is a well-established causal risk factor for atherosclerotic cardiovascular disease (ASCVD), and its reduction is associated with a proportional decrease in cardiovascular events [1]. Contemporary guidelines, including those from the European Society of Cardiology [2], the American Heart Association [3], and the 2025 Taiwan lipid management clinical pathway [4], emphasize a risk-stratified approach to lipid management, with increasingly stringent LDL-C targets for higher-risk populations. In particular, patients with established ASCVD are recommended to achieve LDL-C levels < 55 mg/dL with ≥ 50% reduction from baseline, whereas patients in primary prevention are managed according to overall cardiovascular risk with less stringent but clinically meaningful targets.

However, substantial gaps in LDL-C target attainment persist in real-world practice globally [5] and in Taiwan [6], particularly among high-risk patients. These gaps are often driven by patient characteristics, including cardiovascular risk status, diabetes, age, statin tolerance, and sex, all of which may affect treatment selection, tolerability, and response to lipid-lowering therapy (LLT). The magnitude of LDL-C reduction required to achieve optimal cardiovascular risk reduction may also vary across these groups, further underscoring the importance of subgroup-specific evaluation. Accordingly, understanding treatment effectiveness across clinically relevant patient subgroups is critical to inform individualized lipid management strategies.

Patients with diabetes are a high-risk population characterized by complex metabolic disturbances and elevated cardiovascular risk [7]. Achieving optimal LDL-C control in this population remains challenging due to coexisting comorbidities, treatment complexity, and concerns regarding the metabolic effects of certain LLT [8].

Older adults represent a particularly important population, as advancing age is associated with increased cardiovascular risk and a higher burden of comorbidities. However, LLT in elderly patients is often suboptimal due to concerns regarding polypharmacy, tolerability, and potential adverse effects, leading to persistent unmet needs in achieving LDL-C targets [9].

Statin intolerance represents a major barrier to effective lipid management. Patients who are unable to tolerate statins often receive suboptimal therapy, resulting in inadequate LDL-C reduction and increased cardiovascular risk. This unmet need is further compounded by the potential nocebo effect and limitations of alternative therapies in routine clinical practice [10].

In addition, sex-based differences in lipid management have been increasingly recognized. Women are less likely to receive high-intensity statins and more likely to report statin-associated adverse effects, which may contribute to suboptimal LDL-C control despite comparable or even higher lifetime cardiovascular risk [11].

Bempedoic acid, an oral non-statin LLT that inhibits ATP citrate lyase upstream of HMG-CoA reductase, has demonstrated effective LDL-C reduction with a favorable safety profile in randomized clinical trials [12,13]. Consistent findings have also been observed in the overall population of the CLEAR Taiwan study [14]. However, real-world evidence regarding its effectiveness and safety across clinically relevant patient subgroups remains limited, particularly in Asian populations. Therefore, this subgroup analysis of the CLEAR Taiwan study aimed to evaluate the effectiveness and safety of bempedoic acid across multiple clinically relevant subgroups, including cardiovascular risk category (primary versus secondary prevention), diabetes status, age, statin tolerance, and sex to better inform individualized lipid-lowering strategies in real-world clinical practice.

## MATERIAL AND METHODS

### Study Design

The CLEAR Taiwan study (NCT06925100) is a prospective, pragmatic, phase IV, multicenter, single-arm study conducted at three medical centers in Taiwan [14]. Eligible patients were identified during routine clinical practice according to predefined inclusion and exclusion criteria, and consecutive patients who met these criteria during the study period were invited to participate to minimize selection bias. All patients received bempedoic acid in addition to background LLT for 12 weeks.

The present analyses are prespecified subgroup analyses of the CLEAR Taiwan study, designed to evaluate the effectiveness and safety of bempedoic acid across clinically relevant patient subgroups defined by the following baseline characteristics: (1) cardiovascular risk status (primary versus secondary prevention), (2) diabetes status (with versus without diabetes), (3) statin tolerance (statin-tolerant versus statin-intolerant), (4) age (< 65 versus ≥ 65 years), and (5) sex (male versus female).

The study was conducted in accordance with the principles of the Declaration of Helsinki and the International Council for Harmonization’s Guideline for Good Clinical Practice. Approval was obtained from the ethics committees and institutional review boards of all participating institutions (Linkou Chang Gung Memorial Hospital: 202401853A4; Far Eastern Memorial Hospital: 113314-J; Kaohsiung Medical University Chung-Ho Memorial Hospital: KMUHIRB-F(1)-20240296). Written informed consent was obtained from all participants prior to study enrollment.

### Participants

Adult patients aged ≥ 18 years with a diagnosis of primary hypercholesterolemia or mixed dyslipidemia were eligible if they had inadequately controlled LDL-C levels despite receiving LLT for at least 4 weeks. Inadequate LDL-C control was defined as fasting LDL-C levels ≥ 70 mg/dL in patients with ASCVD (including coronary artery disease, acute coronary syndrome, peripheral artery disease, ischemic stroke, transient ischemic attack, or documented atherosclerotic stenosis of cerebral or carotid arteries) and ≥ 100 mg/dL in patients without ASCVD. Patients with contraindications to bempedoic acid according to the locally approved label were excluded.

### Subgroup Definitions

Primary prevention was defined as the absence of documented ASCVD, whereas secondary prevention included patients with established ASCVD as defined in inclusion criteria above. Statin intolerance was defined as meeting any of the following criteria at screening: (1) use of low- or very-low-dose statin with or without other lipid-lowering therapies; (2) discontinuation of statin therapy; or (3) use of non-statin monotherapy irrespective of prior statin use. Diabetes status was determined based on documented medical history. Age groups were defined as < 65 years and ≥ 65 years, and sex was categorized as male or female.

### Assessments and Endpoints

Lipid parameters, including LDL-C, non–high-density lipoprotein cholesterol (non-HDL-C), total cholesterol, triglycerides, high-density lipoprotein cholesterol (HDL-C), and apolipoprotein B (apoB), as well as high-sensitivity C-reactive protein (hsCRP), were assessed at baseline and week 12. The primary endpoint was the percentage change in LDL-C from baseline to week 12. Secondary endpoints included percentage changes in other lipid parameters and hsCRP. Safety assessments included adverse events, laboratory parameters (including hepatic enzymes, uric acid, and renal function), and treatment adherence.

### Statistical Analysis

These subgroup analyses were performed in patients with available baseline and week 12 data (complete-case analysis), without imputation for missing values. Percentage changes from baseline were calculated for each patient and analyzed as derived variables. Within-group changes from baseline were evaluated using paired t-tests or Wilcoxon signed-rank tests, as appropriate based on data distribution. Between-group comparisons were descriptive in nature and not adjusted for multiplicity. Given the exploratory nature of this subgroup analysis, no formal hypothesis testing for subgroup differences was prespecified, and subgroup comparison results should be interpreted descriptively.

## RESULTS

### Patient Characteristics

A total of 180 patients were enrolled in the CLEAR Taiwan study, of whom 160 (88.9%) completed the 12-week follow-up and were included in the analysis. Baseline characteristics of the overall population are summarized in Table 1. Across prespecified subgroups, patient distributions were as follows: 59 patients (32.8%) in the primary prevention group and 121 (67.2%) in the secondary prevention group; 56 patients (31.1%) with diabetes and 124 (68.9%) without diabetes; 112 patients (62.2%) aged < 65 years and 68 (37.8%) aged ≥ 65 years; 97 (53.9%) statin-tolerant and 83 (46.1%) statin-intolerant patients; and 113 male (62.8%) and 67 female (37.2%) patients (Table 1).

**Table 1.**
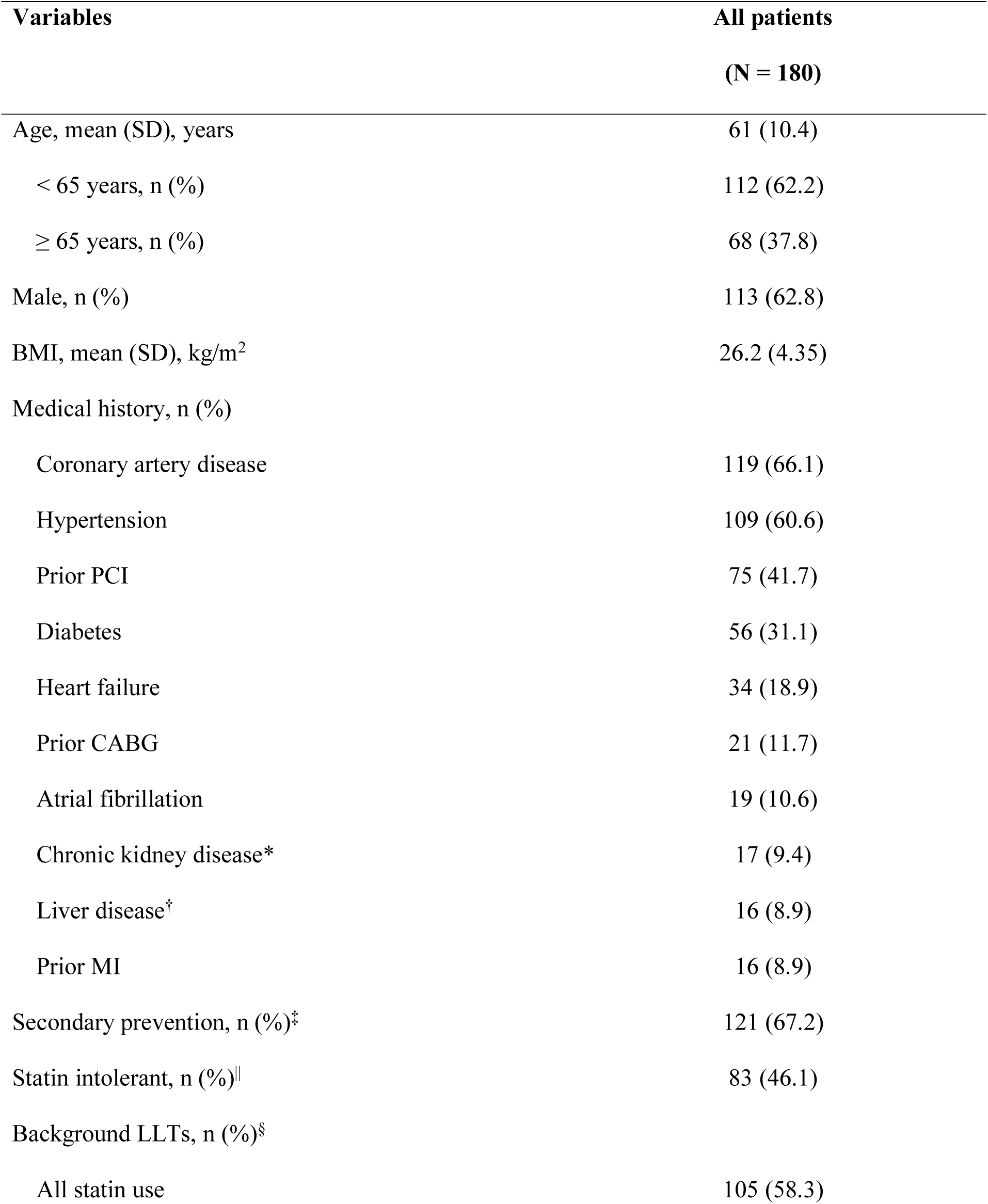
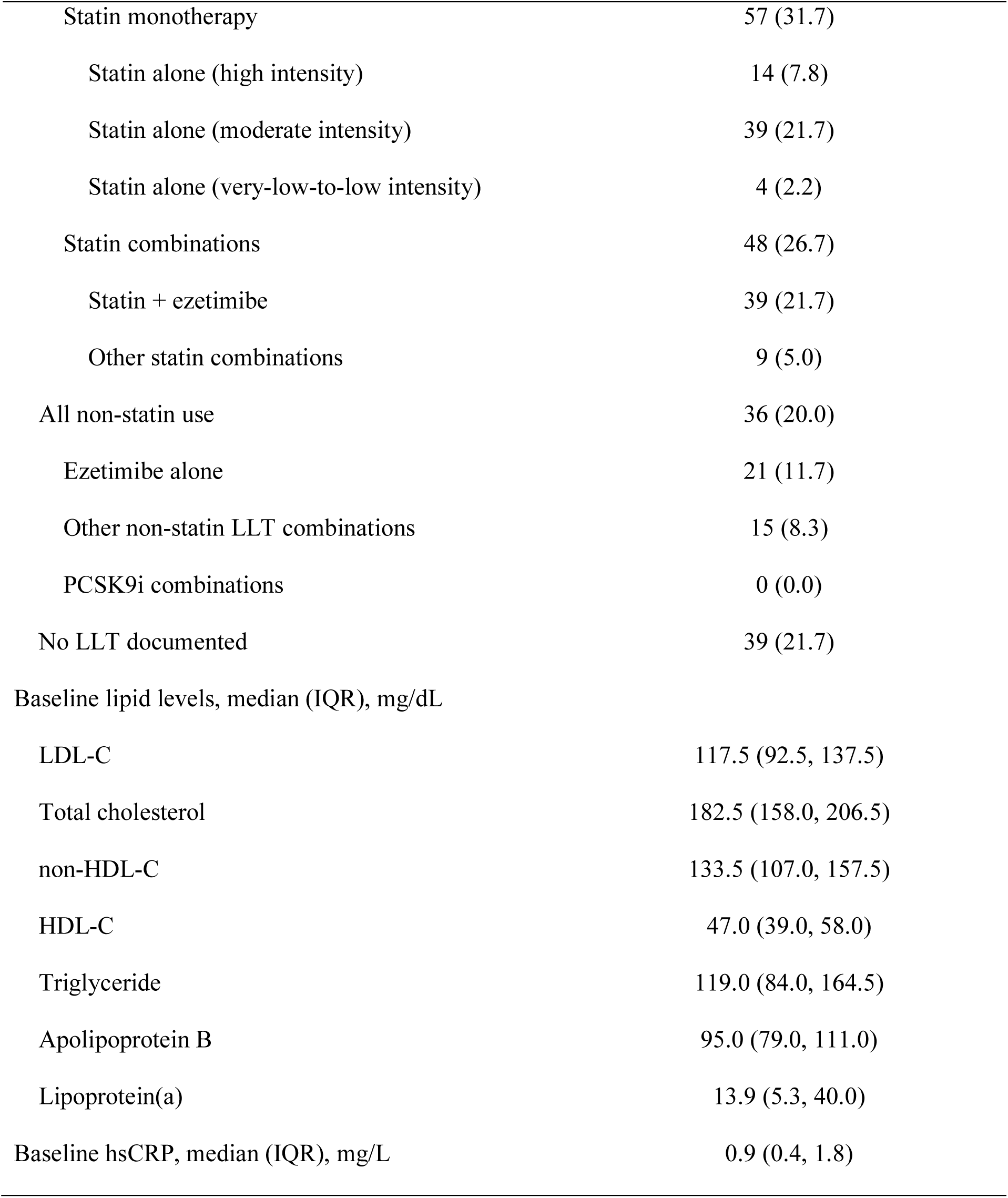
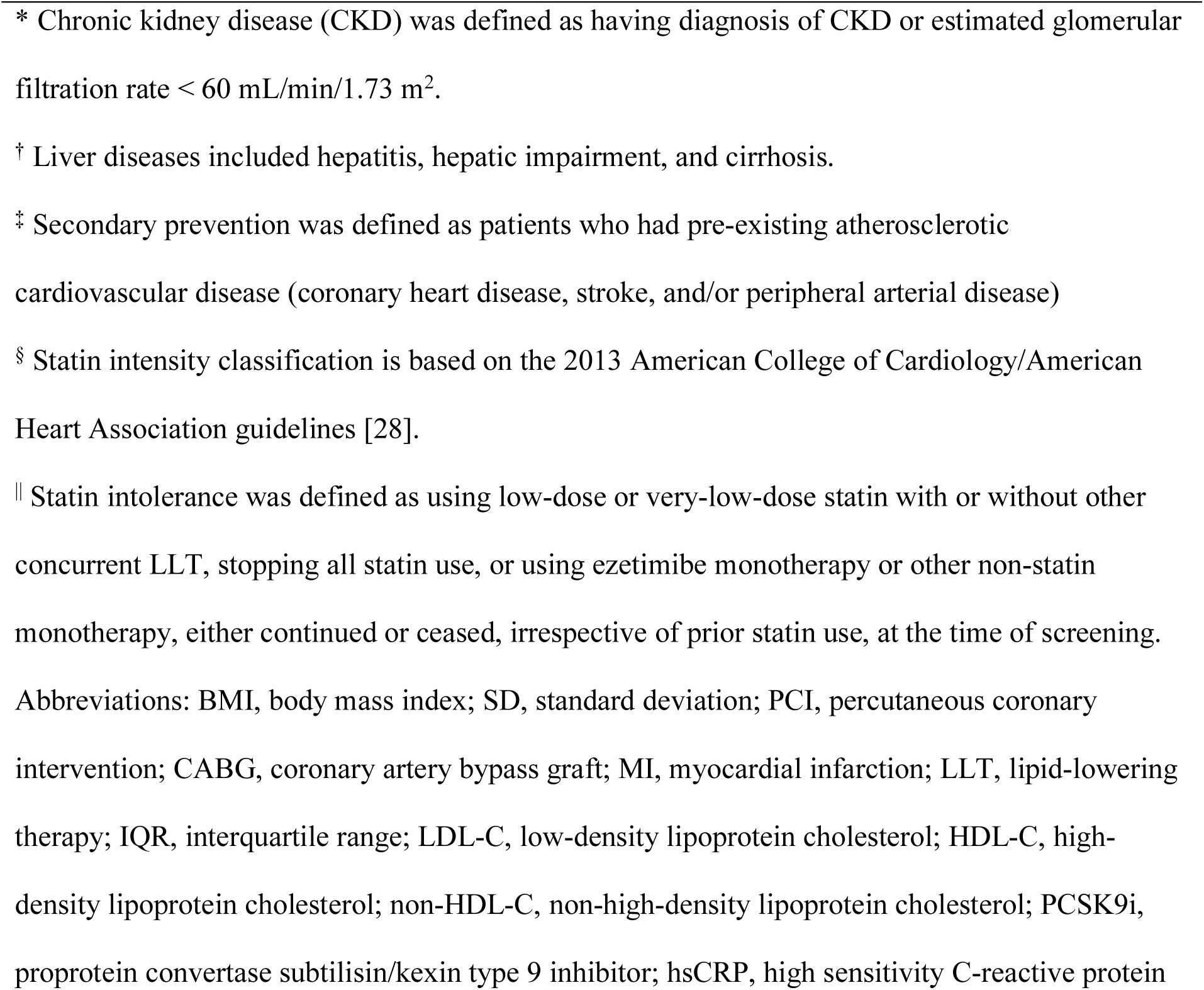
Patient demographics and baseline characteristics.

| **Variables** | **All patients**<br>**(N = 180)** |
| --- | --- |
| Age, mean (SD), years | 61 (10.4) |
| < 65 years, n (%) | 112 (62.2) |
| ≥ 65 years, n (%) | 68 (37.8) |
| Male, n (%) | 113 (62.8) |
| BMI, mean (SD), kg/m<sup>2</sup> | 26.2 (4.35) |
| Medical history, n (%) |  |
| Coronary artery disease | 119 (66.1) |
| Hypertension | 109 (60.6) |
| Prior PCI | 75 (41.7) |
| Diabetes | 56 (31.1) |
| Heart failure | 34 (18.9) |
| Prior CABG | 21 (11.7) |
| Atrial fibrillation | 19 (10.6) |
| Chronic kidney disease* | 17 (9.4) |
| Liver disease<sup>†</sup> | 16 (8.9) |
| Prior MI | 16 (8.9) |
| Secondary prevention, n (%)<sup>‡</sup> | 121 (67.2) |
| Statin intolerant, n (%) | 83 (46.1) |
| Background LLTs, n (%)<sup>§</sup> |  |
| All statin use | 105 (58.3) |
| Statin monotherapy | 57 (31.7) |
| Statin alone (high intensity) | 14 (7.8) |
| Statin alone (moderate intensity) | 39 (21.7) |
| Statin alone (very-low-to-low intensity) | 4 (2.2) |
| Statin combinations | 48 (26.7) |
| Statin + ezetimibe | 39 (21.7) |
| Other statin combinations | 9 (5.0) |
| All non-statin use | 36 (20.0) |
| Ezetimibe alone | 21 (11.7) |
| Other non-statin LLT combinations | 15 (8.3) |
| PCSK9i combinations | 0 (0.0) |
| No LLT documented | 39 (21.7) |
| Baseline lipid levels, median (IQR), mg/dL |  |
| LDL-C | 117.5 (92.5, 137.5) |
| Total cholesterol | 182.5 (158.0, 206.5) |
| non-HDL-C | 133.5 (107.0, 157.5) |
| HDL-C | 47.0 (39.0, 58.0) |
| Triglyceride | 119.0 (84.0, 164.5) |
| Apolipoprotein B | 95.0 (79.0, 111.0) |
| Lipoprotein(a) | 13.9 (5.3, 40.0) |
| Baseline hsCRP, median (IQR), mg/L | 0.9 (0.4, 1.8) |

| Variables | All patients<br>(N = 180) |
| --- | --- |
| * Chronic kidney disease (CKD) was defined as having diagnosis of CKD or estimated glomerular filtration rate < 60 mL/min/1.73 m<sup>2</sup>. |  |
| † Liver diseases included hepatitis, hepatic impairment, and cirrhosis. |  |
| ‡ Secondary prevention was defined as patients who had pre-existing atherosclerotic cardiovascular disease (coronary heart disease, stroke, and/or peripheral arterial disease) |  |
| § Statin intensity classification is based on the 2013 American College of Cardiology/American Heart Association guidelines [28]. |  |
| \|\| Statin intolerance was defined as using low-dose or very-low-dose statin with or without other concurrent LLT, stopping all statin use, or using ezetimibe monotherapy or other non-statin monotherapy, either continued or ceased, irrespective of prior statin use, at the time of screening. |  |
| Abbreviations: BMI, body mass index; SD, standard deviation; PCI, percutaneous coronary intervention; CABG, coronary artery bypass graft; MI, myocardial infarction; LLT, lipid-lowering therapy; IQR, interquartile range; LDL-C, low-density lipoprotein cholesterol; HDL-C, high-density lipoprotein cholesterol; non-HDL-C, non-high-density lipoprotein cholesterol; PCSK9i, proprotein convertase subtilisin/kexin type 9 inhibitor; hsCRP, high sensitivity C-reactive protein |  |

Baseline characteristics varied among the subgroups (Supplementary Table 1 - 5). Patients in the secondary prevention group were older, more often male, and had a higher proportion of diabetes compared to those in the primary prevention group. Statin-intolerant patients were younger, more frequently female, and had a lower proportion of diabetes than statin-tolerant patients. Similarly, female patients had a lower proportion of diabetes compared with male patients. As expected, patients aged ≥ 65 years and those with diabetes had a higher burden of cardiometabolic risk factors.

Differences in background LLT were also observed across subgroups, including variations in statin use, intensity, and combination therapy. Subgroups with a higher proportion of statin-tolerant patients, such as those in the secondary prevention group and male patients, were more likely to receive moderate- to high-intensity statins and combination lipid-lowering therapy. However, statin-intolerant and female patients were more likely to receive lower-intensity or no statin therapy, relying more on non-statin treatments. Similar patterns were observed in younger patients and those without diabetes, who generally used less intensive statin therapy compared to older patients and those with diabetes (Supplementary Table 1 - 5).

### Effectiveness

As previously reported in the overall CLEAR Taiwan population [^14^], bempedoic acid significantly reduced LDL-C by 19% at week 12 and was associated with significant reductions in non-HDL-C, total cholesterol, apoB, and hsCRP. Significant LDL-C reductions were observed within all prespecified subgroups, although the magnitude of reduction varied among groups (Figure 1). Overall, numerically greater LDL-C reductions were observed in primary prevention, younger (< 65 years), statin-intolerant, and female patients, whereas the effects were consistent regardless of diabetes status. Specifically, median LDL-C reductions were 27.3% in the primary prevention subgroup compared to 17.5% in the secondary prevention subgroup. Patients aged < 65 years and ≥ 65 years experienced median LDL-C reductions of 21.7% and 14.5%, respectively. Among statin-tolerant and statin-intolerant patients, median LDL-C reductions were 18.5% and 22.3%, respectively. Female patients showed a numerically greater reduction in LDL-C compared with male patients (25.6% versus 18.0%). In patients with and without diabetes, LDL-C reductions were comparable (20.1% versus 19.0%).

**Figure 1.**
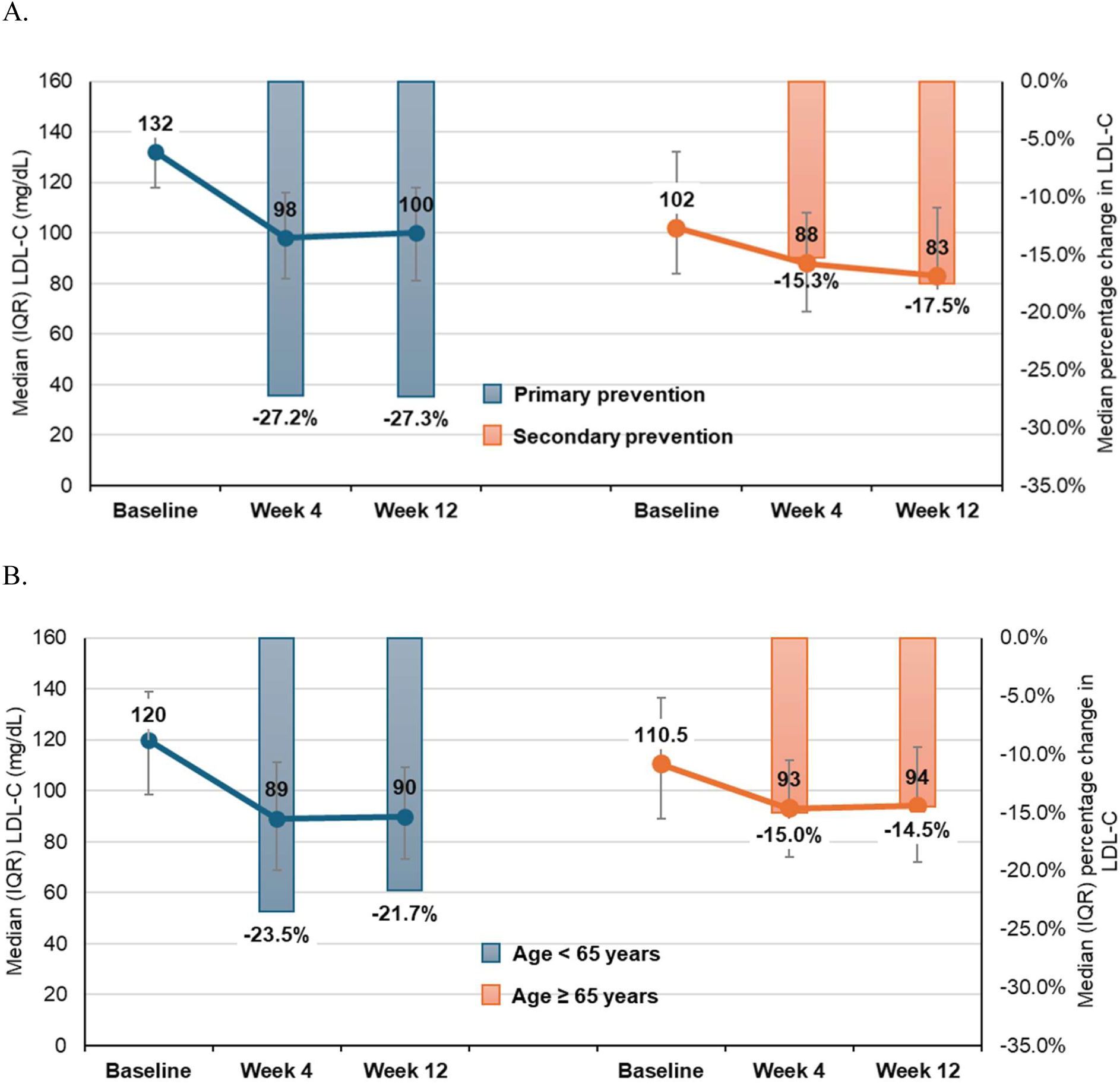
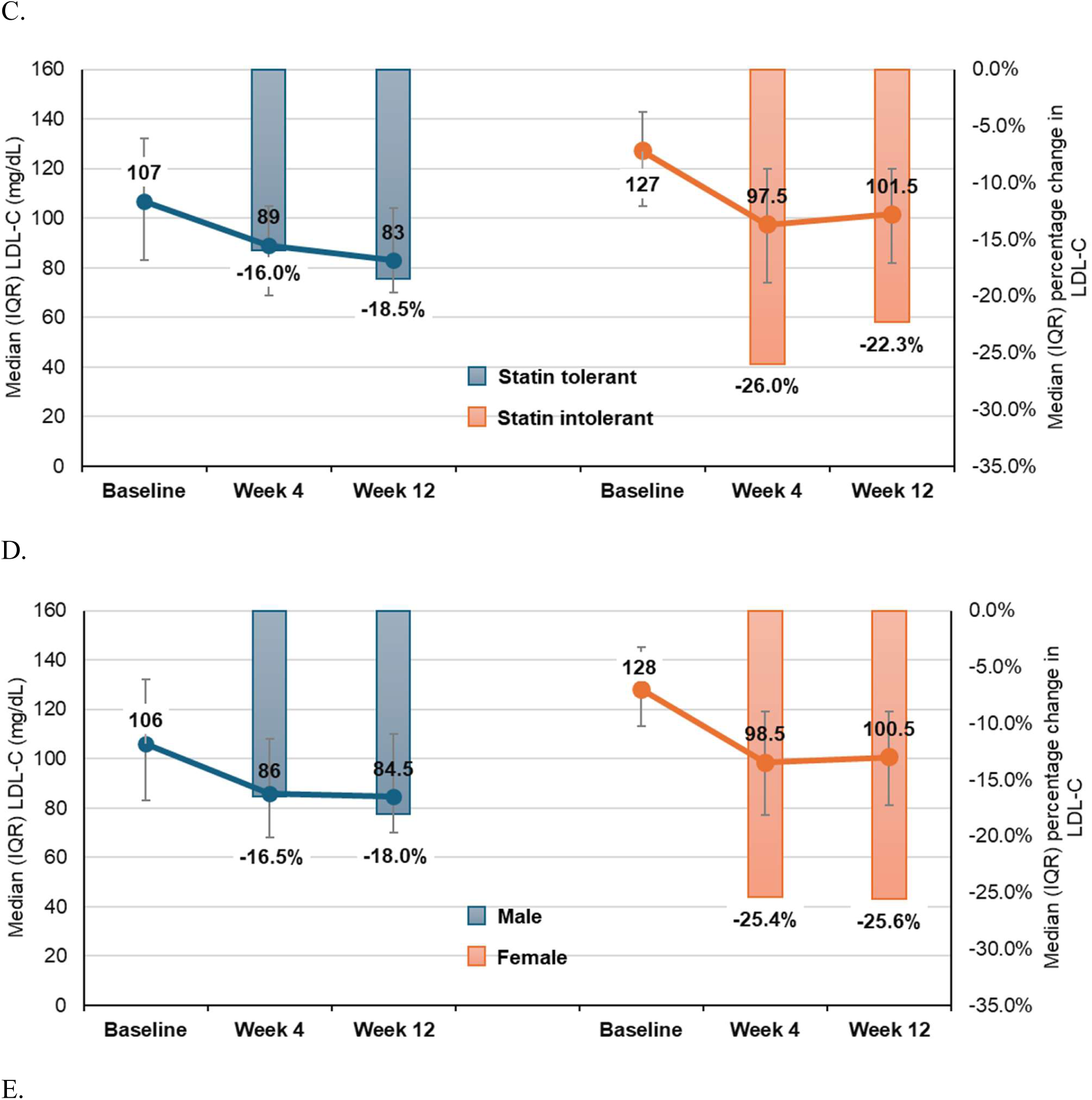
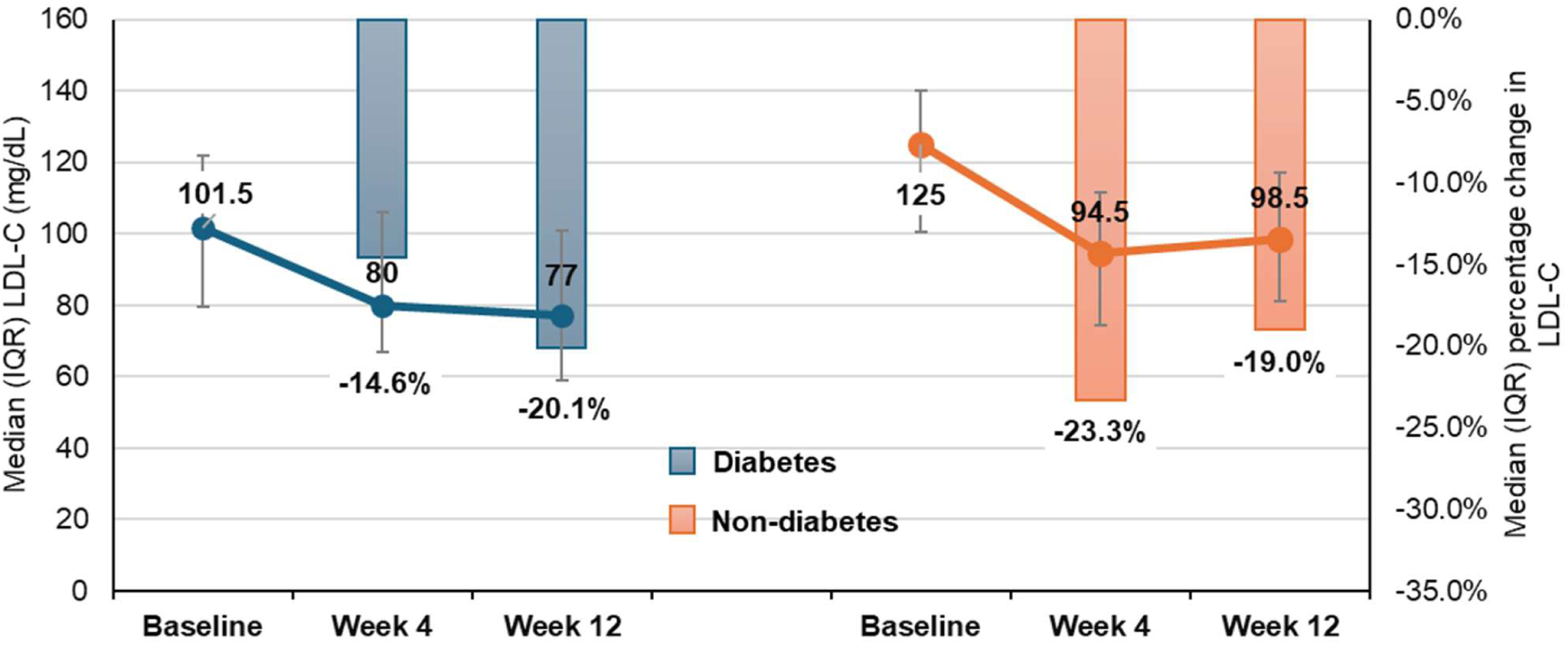
Low-density lipoprotein cholesterol (LDL-C) levels and percentage change from baseline to week 4 and week 12 in LDL-C among subgroups (A) primary prevention versus secondary prevention patients (B) patients aged < 65 years versus ≥ 65 years (C) statin-tolerant versus statin-intolerant patients (D) male versus female patients (E) patients with and without diabetes. Data are presented as median with interquartile range (IQR).

Reductions in non-HDL-C, total cholesterol, and apoB generally mirrored those observed in LDL-C across all prespecified subgroups (Supplementary Table 6 - 10). Significant reductions in hsCRP were also observed within all subgroups, with similar magnitudes between primary and secondary prevention groups (−34.0% versus -33.3%), statin-tolerant and statin-intolerant patients (−33.5% versus -34.1%), as well as male and female patients (both -33.6%). Notably, certain subgroups demonstrated numerically greater reductions, including patients aged < 65 years (−36.2% versus -16.7% in patients ≥ 65 years) and patients without diabetes (−33.9% versus -17.9% in patients with diabetes) (Supplementary Table 6 - 10).

With respect to glycemic parameters, a small reduction in HbA1c (−0.1%) was observed among patients with diabetes, reaching nominal statistical significance (*p* < 0.05). No changes were observed in patients without diabetes (Supplementary Table 7).

### Safety

Safety outcomes were generally consistent across all subgroups, in line with those observed in the overall CLEAR Taiwan population [14]. The incidence of adverse events was low and comparable between groups. Most events were mild and manageable, with no new safety signals identified.

Elevations in liver enzymes were observed in a small proportion of patients. The incidence was numerically higher in younger patients (< 65 years), those with diabetes, secondary prevention group, and statin-intolerant patients; however, most cases were mild and did not lead to treatment discontinuation. Clinically significant elevations (≥ 3 × upper limit of normal) were infrequent in all subgroups (Table 2).

**Table 2.**
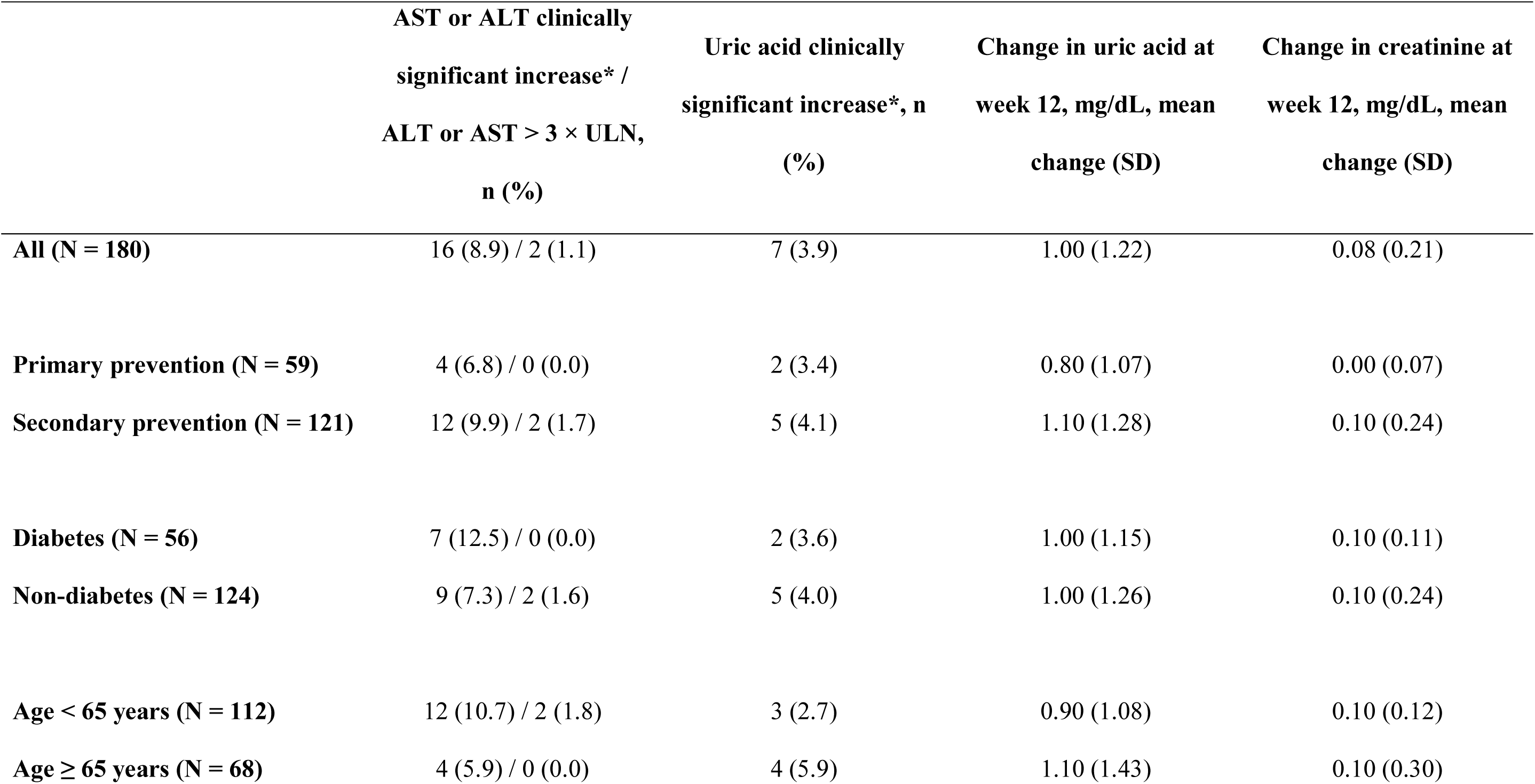
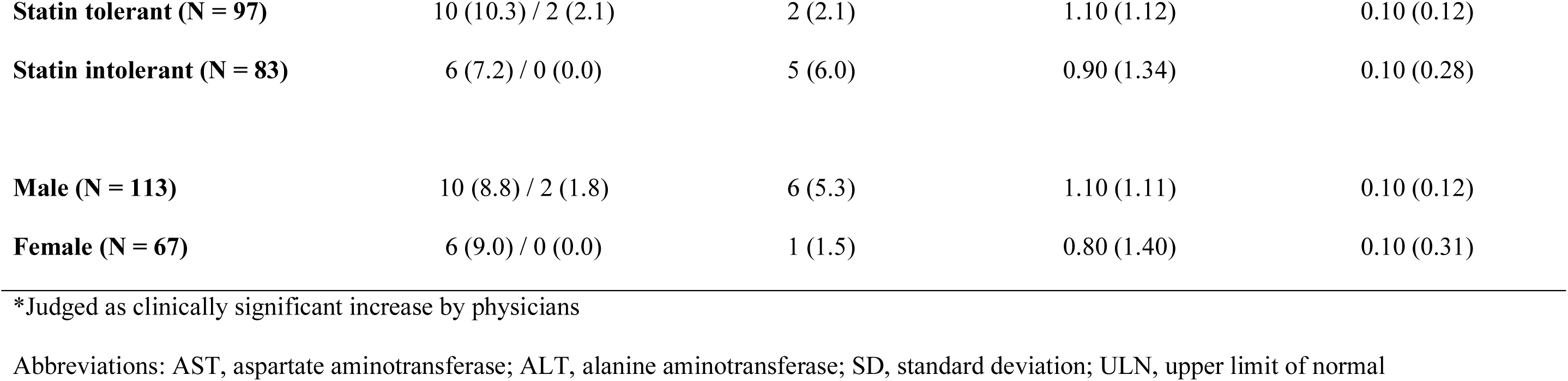
Changes in liver and kidney function tests and related adverse events.

|  | AST or ALT clinically<br>significant increase* /<br>ALT or AST > 3 × ULN,<br>n (%) | Uric acid clinically<br>significant increase*, n<br>(%) | Change in uric acid at<br>week 12, mg/dL, mean<br>change (SD) | Change in creatinine at<br>week 12, mg/dL, mean<br>change (SD) |
| --- | --- | --- | --- | --- |
| **All (N = 180)** | 16 (8.9) / 2 (1.1) | 7 (3.9) | 1.00 (1.22) | 0.08 (0.21) |
| **Primary prevention (N = 59)** | 4 (6.8) / 0 (0.0) | 2 (3.4) | 0.80 (1.07) | 0.00 (0.07) |
| **Secondary prevention (N = 121)** | 12 (9.9) / 2 (1.7) | 5 (4.1) | 1.10 (1.28) | 0.10 (0.24) |
| **Diabetes (N = 56)** | 7 (12.5) / 0 (0.0) | 2 (3.6) | 1.00 (1.15) | 0.10 (0.11) |
| **Non-diabetes (N = 124)** | 9 (7.3) / 2 (1.6) | 5 (4.0) | 1.00 (1.26) | 0.10 (0.24) |
| **Age < 65 years (N = 112)** | 12 (10.7) / 2 (1.8) | 3 (2.7) | 0.90 (1.08) | 0.10 (0.12) |
| **Age ≥ 65 years (N = 68)** | 4 (5.9) / 0 (0.0) | 4 (5.9) | 1.10 (1.43) | 0.10 (0.30) |
| **Statin tolerant (N = 97)** | 10 (10.3) / 2 (2.1) | 2 (2.1) | 1.10 (1.12) | 0.10 (0.12) |
| **Statin intolerant (N = 83)** | 6 (7.2) / 0 (0.0) | 5 (6.0) | 0.90 (1.34) | 0.10 (0.28) |
| **Male (N = 113)** | 10 (8.8) / 2 (1.8) | 6 (5.3) | 1.10 (1.11) | 0.10 (0.12) |
| **Female (N = 67)** | 6 (9.0) / 0 (0.0) | 1 (1.5) | 0.80 (1.40) | 0.10 (0.31) |

\*Judged as clinically significant increase by physicians
Abbreviations: AST, aspartate aminotransferase; ALT, alanine aminotransferase; SD, standard deviation; ULN, upper limit of normal

Clinically significant increases in uric acid were observed in a subset of patients. The incidence was numerically higher among older (≥ 65 years), statin-intolerant, and male patients, whereas comparable incidence was observed between primary and secondary prevention groups and by diabetes status (Table 2). Despite these differences, the magnitude of change in uric acid levels was generally small, and no clinically meaningful renal impairment or gout-related complications were reported. Changes in creatinine levels were minimal and comparable between groups (Table 2). No clinically meaningful deterioration in renal function was observed. Overall, the safety profile of bempedoic acid was consistent regardless of cardiovascular risk status, age, sex, diabetes status, or statin tolerance, and was in line with previous studies and the locally approved label.

## DISCUSSION

In these prespecified subgroup analyses of the CLEAR Taiwan study, bempedoic acid demonstrated consistent lipid-lowering efficacy and a favorable safety profile across all clinically relevant patient subgroups, including cardiovascular risk status, age, sex, diabetes status, and statin tolerance. Although the magnitude of LDL-C reduction varied among subgroups, the overall treatment effect was preserved, with numerically greater reductions observed in primary prevention, statin-intolerant, younger, and female patients, while comparable effects were observed across diabetes status. These findings are generally consistent with previous analyses of bempedoic acid trials, which have demonstrated greater lipid-lowering effects in statin-intolerant patients [15,16] and women [17], while showing comparable efficacy across glycemic strata [18]. The observed variability across subgroups may be partly attributable to differences in baseline LDL-C levels, background LLT intensity, and statin exposure rather than intrinsic differences in treatment responsiveness. In addition to LDL-C lowering, improvements in other atherogenic lipid parameters and hsCRP were consistently observed across subgroups, supporting the broad therapeutic effect of bempedoic acid. The safety profile was similarly consistent, with a low incidence of adverse events and no new safety signals identified, in line with those reported in the overall CLEAR Taiwan population [14]. Mild elevations in liver enzymes and uric acid were observed in a small proportion of patients, with numerical differences between groups and no clinically meaningful consequences. Overall, these findings highlight the robustness and generalizability of bempedoic acid across clinically diverse patient populations.

Bempedoic acid demonstrated clinically meaningful LDL-C reductions in both primary and secondary prevention populations, supporting its role across the continuum of cardiovascular risk and highlighting its potential utility in earlier stages of lipid management. Notably, numerically greater LDL-C reductions were observed in the primary prevention group, which may be partly explained by differences in baseline LDL-C level and LLT intensity, as patients in the secondary prevention population were more likely to receive background moderate- to high-intensity statins and combination LLT, thereby limiting the incremental LDL-C-lowering effect of add-on bempedoic acid. However, as a non-statin lipid-lowering agent, bempedoic acid may be more commonly used in the secondary preventive setting to achieve more stringent guideline-directed risk factor control. These findings support the consideration of bempedoic acid as a practical option not only for treatment intensification in secondary prevention, but also for earlier intervention in high-risk primary prevention patients. Early initiation of bempedoic acid in such populations may facilitate timely achievement of LDL-C targets and potentially reduce long-term cardiovascular risk.

Lipid-lowering efficacy was maintained across age groups, together with favorable safety profiles, supporting the use of bempedoic acid in both younger and older patients. In older patients, for whom treatment intensification is often limited by concerns related to tolerability, comorbidities, and polypharmacy, the observed efficacy and favorable safety profile are of clinical relevance. In the present study, numerically greater LDL-C reductions were observed in patients aged < 65 years compared with those aged ≥ 65 years. This difference may be attributable to more frequent use of intensive background LLT in older patients, which may attenuate the relative treatment effect of bempedoic acid. Prior studies have demonstrated comparable efficacy of bempedoic acid across age groups. The pooled phase 3 trials reported that bempedoic acid reduced LDL-C by approximately 18% to 28% depending on background therapy. Subgroup analyses indicate that the magnitude of LDL-C reduction and overall safety profile did not materially differ by age [19]. Similarly, emerging real-world evidence from studies in elderly populations has consistently demonstrated comparable reductions in LDL-C and other lipid parameters across age groups with bempedoic acid [20,21,22]. In addition, these studies have shown no clinically meaningful differences in renal or hepatic safety profiles between younger and older patients, supporting the use of bempedoic acid as a well-tolerated add-on therapy in elderly individuals. These findings are aligned with the results of the present study and further reinforce its applicability in older patients in routine clinical practice.

The significant LDL-C reductions observed in both statin-tolerant and statin-intolerant patients underscore the clinical utility of bempedoic acid across a broad spectrum of treatment scenarios, particularly in statin-intolerant patients, a group that is frequently undertreated in real-world settings. In this study, statin-intolerant patients exhibited numerically greater LDL-C reductions compared with statin-tolerant patients. This finding is likely attributable to lower baseline statin use and treatment intensity, allowing for a greater relative reduction with the addition of bempedoic acid. This observation is consistent with findings from prior phase 3 trials, in which greater absolute reductions in LDL-C were reported in statin-intolerant patients (33.6 - 39.3 mg/dL [15,16]) compared with those receiving maximally tolerated statin therapy (19.2 - 21.8 mg/dL [23,24]). The clinical relevance of these findings is further supported by the recent 2026 Taiwan Society of Lipids and Atherosclerosis consensus statement, which emphasizes the need for effective non-statin therapies in patients receiving suboptimally tolerable statins [25]. In this context, bempedoic acid represents a practical and well-tolerated therapeutic option that may help bridge existing treatment gaps.

In the present study, female patients exhibited numerically greater LDL-C reductions compared with male patients. This difference may be partly explained by variations in baseline characteristics and treatment patterns, as female patients were more likely to be statin-intolerant and to receive lower-intensity lipid-lowering therapy, potentially allowing for a greater relative treatment effect. These findings are directionally consistent with prior observations that women are less likely to receive high-intensity statin therapy and more likely to experience statin-associated adverse effects [11]. Similarly, pooled phase 3 analyses have reported greater LDL-C reductions in women, with consistent improvements in lipid and inflammatory biomarkers in both sexes [17]. However, in the prespecified sex analysis of CLEAR (Cholesterol Lowering via Bempedoic acid, an ACL-Inhibiting Regimen) Outcomes trial, LDL-C reduction, cardiovascular risk reduction, and adverse event rates were similar between females and males, with no significant treatment-by-sex interaction [26]. Collectively, these findings suggest that the observed differences are more likely driven by baseline treatment patterns rather than intrinsic sex-related differences in drug response. Overall, bempedoic acid represents an effective and well-tolerated therapeutic option in both sexes and may help address existing treatment gaps in female patients.

LDL-C reductions were similar in patients with and without diabetes, suggesting that glycemic status did not materially impact the lipid-lowering response to bempedoic acid. Notably, the absence of adverse glycemic effects, together with the small reduction in HbA1c among patients with diabetes (−0.1%, with nominal *p* < 0.05), is of clinical importance, given concerns regarding the metabolic effects of certain LLT [8]. These findings are consistent with prior studies demonstrating that bempedoic acid maintains lipid-lowering efficacy without adversely affecting glycemic control in patients with diabetes. In the prespecified diabetes analysis of CLEAR Outcomes trial, patients with diabetes experienced statistically significant cardiovascular benefit, with no increased risk of new-onset diabetes or worsening glycemic control was observed in those without diabetes [27]. Similarly, the pooled phase 3 analyses reported consistent LDL-C reductions across glycemic strata, with no clinically meaningful changes in fasting glucose, HbA1c, or incidence of new-onset diabetes over one year [18]. Collectively, these findings support the use of bempedoic acid as a metabolically neutral LLT in patients with coexisting diabetes.

In the present study, numerically greater reductions in hsCRP were observed in younger patients (< 65 years) and those without diabetes, which may reflect variations in baseline inflammatory status, metabolic profiles, or sample size-related variability. Notably, the observed pattern in patients without diabetes differs from prior analyses, in which greater hsCRP reductions were reported in patients with diabetes [18]. Evidence regarding age-related differences in hsCRP response to bempedoic acid remains limited, and the findings should be interpreted with caution given the relatively small sample size, the inherent variability of hsCRP, and the absence of adjustment for potential confounders. Further studies with larger sample sizes are warranted to better characterize the anti-inflammatory effects of bempedoic acid across age and glycemic subgroups.

Several limitations of this study should be acknowledged. First, the single-arm design without a comparator group limits the ability to draw causal inferences regarding treatment effects. Second, the relatively short duration of follow-up (12 weeks) precludes assessment of long-term efficacy and safety outcomes. In addition, variability in background lipid-lowering therapy across subgroups, including differences in statin use and intensity, was not adjusted for and may have influenced the observed treatment responses. Data on lifestyle factors, dietary habits, and adherence to background therapies were not systematically collected, which may also contribute to inter-individual variability. Finally, the present study was not powered to formally evaluate treatment-effect heterogeneity, and sample sizes within individual subgroups were relatively limited. Therefore, formal interaction analyses were not performed. Subgroup findings should be considered exploratory and hypothesis-generating. These findings should be interpreted descriptively.

## CONCLUSION

The subgroup findings from the CLEAR Taiwan study demonstrated consistent efficacy and safety of bempedoic acid across a range of clinically relevant patient populations, including those commonly encountered in routine practice who present therapeutic challenges. These results support the role of bempedoic acid as a flexible treatment option to address residual gaps in lipid management and may help inform more individualized lipid-lowering strategies in clinical practice.

## Data Availability

Data collected within this study will be made available to researchers after contacting the corresponding author and upon approval by the study sponsor. The researcher should describe the required data and purposes in a proposal. A data sharing agreement will be set up by the study sponsor. All data provided are anonymized to respect the privacy of patients who have participated in the trial in line with applicable laws and regulations.

## ACKNOWLEDGMENTS

The authors would like to express their appreciation to all sub-investigators and participating site study teams for their collaboration and dedication to this multicenter study.

## FUNDING SOURCES

This work was supported by Daiichi Sankyo Taiwan, Ltd.

## AUTHOR CONTRIBUTIONS

IC Hsieh, YW Wu, TH Lin, CY Huang, and WC Yang developed the study concept and design. IC Hsieh, YW Wu, TH Lin, DY Chen, CS Chu, YY Chang, BH Tzeng, TC Huang, HH Lin, WP Chuang, CC Huang, JK Yeh, CY Chu, and MY Ho acquired the data. IC Hsieh, YW Wu, TH Lin, and CY Huang analyzed the data. CY Huang wrote the first draft of the manuscript. IC Hsieh, YW Wu, and TH Lin provided critical revision of the manuscript for important intellectual content. DY Chen, CS Chu, YY Chang, BH Tzeng, TC Huang, HH Lin, WP Chuang, CC Huang, JK Yeh, CY Chu, MY Ho, and WC Yang jointly developed the structure and arguments of the paper. All authors reviewed and approved the final manuscript.

## DISCLOSURES AND CONFLICTS OF INTEREST

The authors have read and confirmed their agreement with the International Committee of Medical Journal Editors authorship and conflict of interest criteria. The authors have also confirmed that this article is unique and not under consideration by nor published in any other publication and that they have permission from rights holders to reproduce any copyrighted material. CY Huang and WC Yang are employees of Daiichi Sankyo Taiwan Ltd. All other authors have declared no conflicts of interest.

## SUPPLEMENTAL MATERIAL

Tables S1–S10

## Non-standard Abbreviations and Acronyms

LLT: lipid-lowering therapy

## Notes

### Competing Interest Statement

Ching-Ya Huang and Wei-Chen Yang are employees of Daiichi Sankyo Taiwan Ltd.

### Clinical Trial

URL: https://clinicaltrials.gov/study/NCT06925100; Unique identifier: NCT06925100

### Author Declarations

Approval was obtained from the ethics committees and institutional review boards of all participating institutions (Linkou Chang Gung Memorial Hospital: 202401853A4; Far Eastern Memorial Hospital: 113314-J; Kaohsiung Medical University Chung-Ho Memorial Hospital: KMUHIRB-F(1)-20240296). Written informed consent was obtained from all participants prior to study enrollment.

